# Strategies to Reduce Hyperglycemia-Related Anxiety in Elite Athletes with Type 1 Diabetes: A Qualitative Analysis

**DOI:** 10.1101/2024.10.19.24315806

**Authors:** Alexandra Katz, Aidan Shulkin, Marc-André Fortier, Jane E. Yardley, Jessica Kichler, Asmaa Housni, Meryem K. Talbo, Rémi Rabasa-Lhoret, Anne-Sophie Brazeau

## Abstract

**Objective:** Managing blood glucose levels is challenging for elite athletes with type 1 diabetes (T1D) as competition can cause unpredictable fluctuations. Hyperglycemia-related anxiety (HRA) likely affects performance and diabetes management, but research is limited. This study investigates current strategies employed to mitigate HRA during competition and the development of alternative approaches.

**Research Design and Methods:** Elite athletes with TID, aged ≥14 who self-reported HRA during competition were recruited. Elite athletes were defined as individuals exercising >10 hours per week whose athletic performance has achieved the highest competition level. 60 to 90-minute virtual semi-structured interviews were analyzed using an Interpretative Phenomenological Analysis.

**Results:** Ten elite athletes with T1D (average age 25 ± 3 years; T1D duration 12 ± 8 years; # of competitions per year 27 ± 19; training time per week 12 ± 6 hours) reported the strategies they currently use to mitigate HRA. These strategies include managing insulin and nutrition intake, embracing social support networks, using technology, practicing relaxation techniques, establishing routines, performing pre-competition aerobic exercise, and maintaining adequate sleep hygiene. Several additional approaches that could be implemented were identified including establishing targeted support networks, developing peer-reviewed resources on HRA, ensuring support teams have sufficient tools, and improving existing technology.

**Conclusions:** Elite athletes with T1D use physiological and psychological strategies to mitigate HRA during competition. This finding highlights the need for increased support and education for these athletes, and advancements in technology. Targeted strategies and personalized approaches are also needed to optimize performance and diabetes management in this population.

## Introduction

Individuals with type 1 diabetes (T1D) compete at the highest level of their sports, including winning Olympic gold medals and becoming professional athletes [1]. However, participating in physical activity (PA) poses considerable challenges for athletes with T1D, primarily due to its multifaceted influence on glycemia [2–5]. T1D is an autoimmune disorder in which the insulin-producing beta cells of the pancreas are destroyed, leading to a significant reduction or complete cessation of insulin production. Incorrect management of blood glucose (BG) modifiers, including insulin, pre-competition meal composition, exercise duration and intensity, stress levels, and sleep quality can lead to dangerous fluctuations in BG levels which can result in hyper- or hypoglycemia [6, 7].

Most people with T1D prioritize avoiding hypoglycemia during PA as it can compromise performance [16] due to lack of concentration, headaches, dizziness, and confusion [17]. Conversely, athletes may also worry about hyperglycemia, which can also reduce performance through debilitating symptoms such as shortness of breath, thirst, slower reaction time, and blurred vision [3, 11].

Fear of hypoglycemia during PA is well documented [9], however, research on hyperglycemia-related anxiety (HRA) is limited. Hyperglycemia is often considered a chronic condition, with its acute symptoms typically causing minimal disruption to daily life [10]. However, any element that hinders competition performance in elite athletes is a significant concern [11]. Various factors, including competition-associated stress may cause hyperglycemia [2, 12, 13]. A position statement from the *National Athletic Trainers’ Association* [14] acknowledges that high anticipatory stress can elevate counterregulatory hormones, such as adrenaline and cortisol, potentially increasing BG before and during competition [9]. This specifically affects elite athletes, as they face increased amounts of performance-related stress and anxiety [8].

Current recommendations for diabetes management are not adapted to the specific physiological and psychological demands of competition [13, 15]. For example, athletes who expect a BG rise before competition are more prone to taking preventive measures (e.g., injecting more insulin, and limiting pre-PA carbohydrate intake) [2].

A recent case study suggests a need for tailored treatment programs for elite athletes with T1D [11], but there is limited information available in the scientific literature and none exploring HRA. Therefore, this study aims to understand the impact that HRA has on competition and diabetes management among elite athletes with T1D. Additionally, it explores the strategies used by these athletes to prevent HRA during competition and maps out additional approaches that they can use to optimize BG management.

## Methods

### Study Design, Participants & Recruitment

This cross-sectional qualitative analysis was conducted following the Consolidated Criteria for Reporting Qualitative Research checklist [18]. Ethics approval was obtained by the Centre Hospitalier de l’Université de Montréal ethics committee (2024-11786 (23.156)). Individuals were eligible to participate in this study if they were elite athletes ≥14 years old, had a T1D diagnosis for ≥1 year, self-reported HRA during competition, used intensive insulin therapy, and spoke English or French. Participants were excluded if they had a diagnosis of Generalized Anxiety Disorder (GAD) or a score of >10 on the GAD 7-item scale [19]. Elite athletes were defined as individuals exercising >10 h/week whose athletic performance had achieved the highest competition level (definition based on [20]). HRA was defined as the feelings of worry, fear, or distress that individuals with T1D had in relation to hyperglycemia. Recruitment was conducted via word-of-mouth, at the Montreal Clinical Research Institute (IRCM) clinic, and by internet postings (e.g., newsletters, websites, social media, etc.). Potential participants were screened for eligibility, consented to participation, filled out a demographic questionnaire, and registered for an interview via the Research Electronic Data Capture (REDCap) tool [21].

### Interviews

Semi-structured individual interviews were scheduled for 60-90 minutes on Microsoft Teams and were conducted in English or French. To enhance the understanding of participant perspectives and acknowledge the potential impact of the researcher’s positionality, the research team employed bracketing, defined as the systematic suspension of preconceptions and biases during data collection and analysis [22]. A research team member followed an interview discussion guide (Figure 1) to direct the open-ended conversation using five probing and additional prompting questions derived from the *Hyperglycemia Avoidance Scale* [23]. The guide was reviewed by a multidisciplinary team including dietitians, an endocrinologist, a clinical psychologist, a kinesiologist, and a patient partner. Before the interview, participants were given the probing questions to reflect on. Additional resources were available for mental health support if necessary.

**Figure 1.**
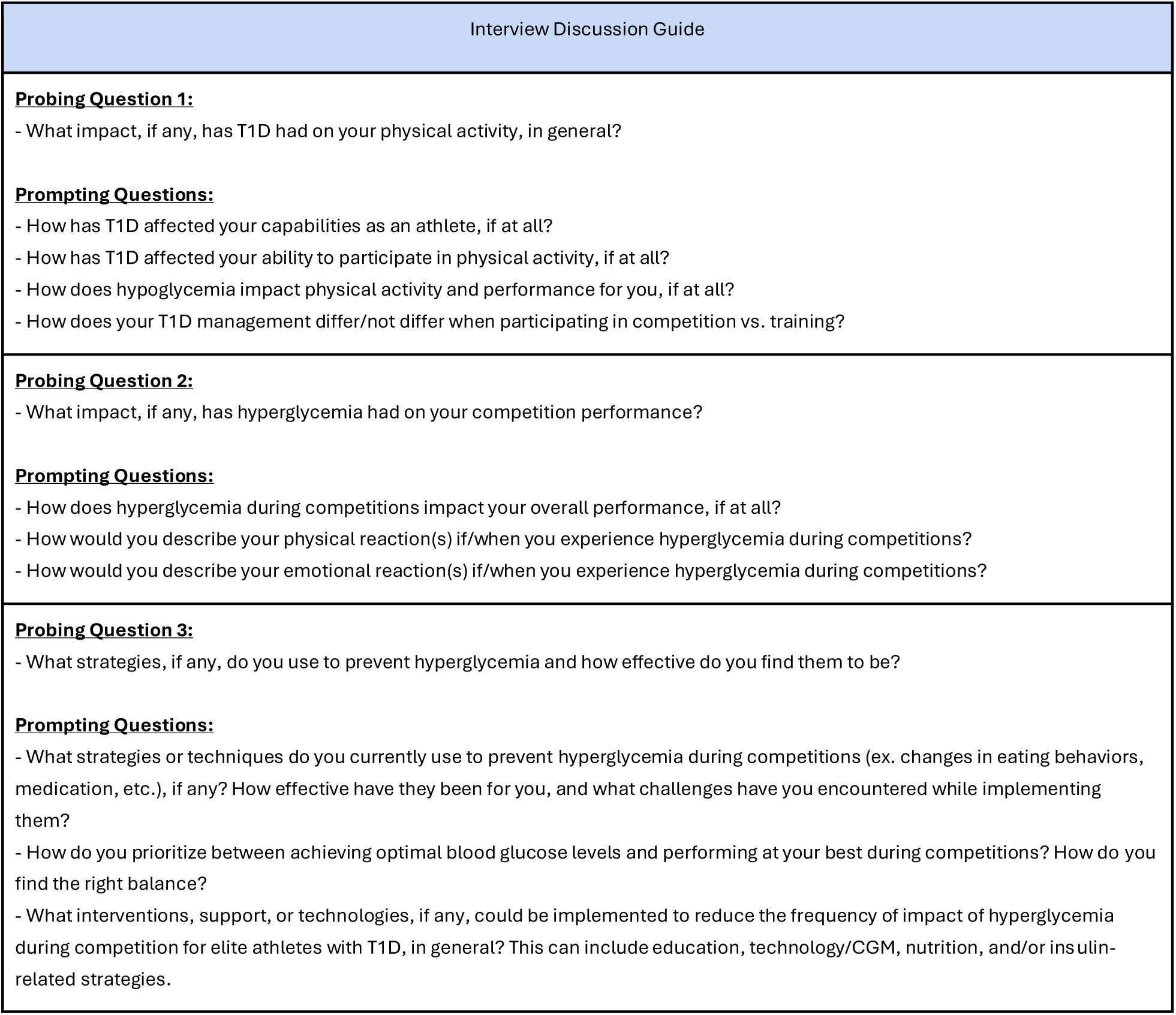
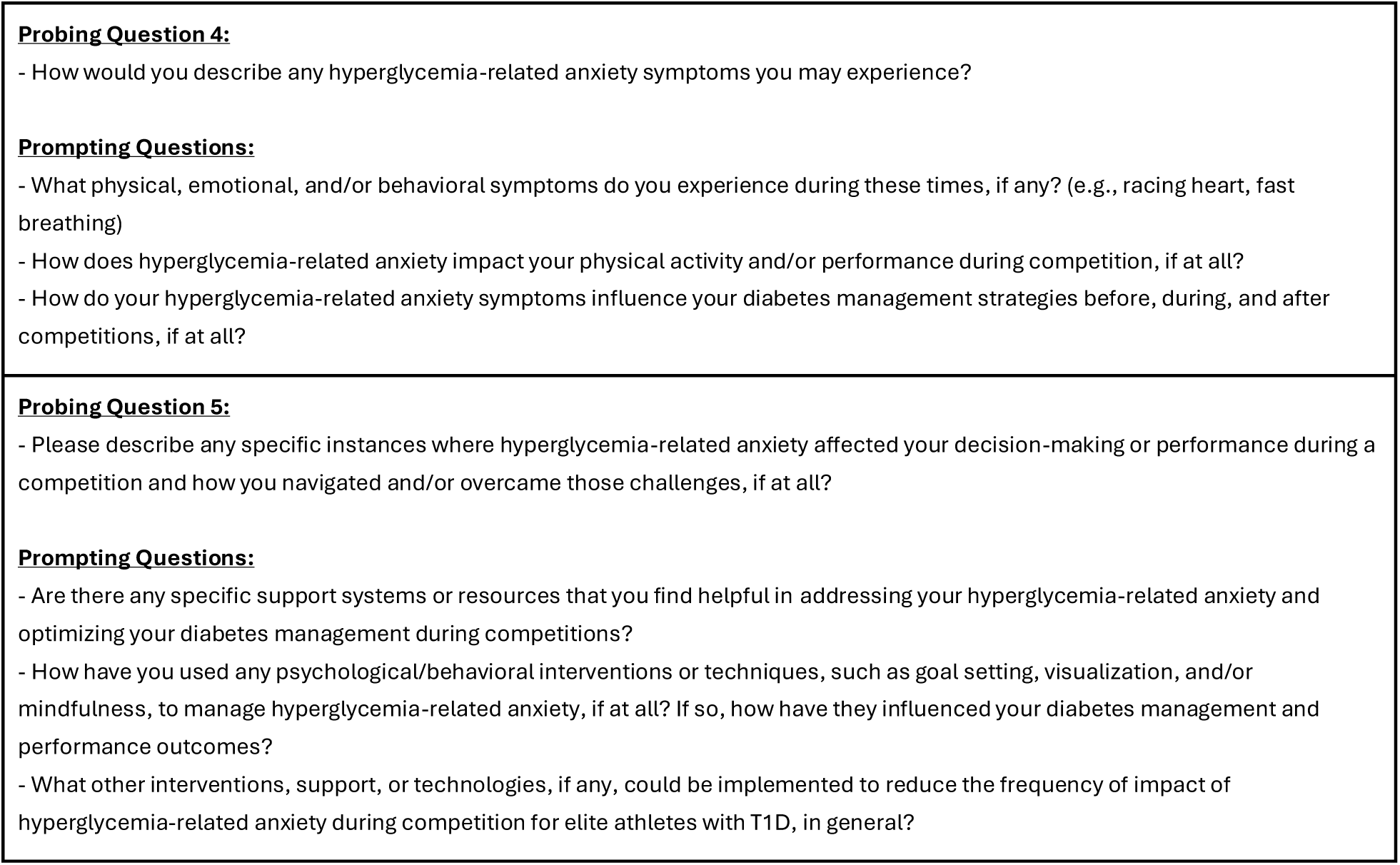
Interview Discussion Guide

### Transcript and Analysis

Interviews were recorded and transcribed using Microsoft Teams. A research team member (MA.F.) verified and anonymized the transcriptions. Recordings were deleted to preserve confidentiality. French meeting transcriptions were translated to English using a forward-backwards translation method.

Transcriptions were uploaded to MAXQDA (version 24, VERBI GmbH, Berlin, Germany) [24]. The beta AI Assist function was used to generate preliminary codes. Using an Interpretive Phenomenological Analysis approach, two independent researchers (A.K. and A.S.) reviewed and refined the codes to create the codebook, which was applied to the entire dataset. Overarching concepts, relationships, or sequences among the themes were identified and analyzed. A detailed essence description of participants’ experiences and highlighted main themes was created. Study participants verified the essence description to complete the member-checking process. Once validated, revisions were made based on feedback. Participants were provided with a 50$ CAD electronic Amazon gift card or a donation in their name to a diabetes-specific charity of their choice.

## Results

Ten elite athletes (5 males, and 5 females) were interviewed from November to December 2023. Participants had an average age of 25 ± 3 years, had a T1D duration of 12 ± 8 years, participated in 27 ± 19 competitions per year, and performed 12 ± 6 hours of training per week. Four participants were tier 3, and six participants were tier 4 athletes (tiers defined by [24]). Four participants used continuous subcutaneous insulin infusions (CSII) and six participants used multiple daily injections. All athletes used real-time continuous glucose monitoring (rtCGM) and its alerts/alarm feature in the last year. They originated from Canada, the United States, or the United Kingdom and self-reported as Caucasian. The athletes engaged in various sports, including speed skating, cross-country/track running, ice hockey, rugby, volleyball, basketball, and baseball.

### Hyperglycemia in Elite Athletes with T1D

#### Hyperglycemia Symptoms

Participants reported experiencing a variety of hyperglycemia symptoms during both training and competition. These included irritability, muscle cramps, impatience, agitation, attention deficiency, excessive thirst, blurred vision, delayed reaction time, frequent urination, fatigue, abdominal pain, and shortness of breath. Elite athletes highlighted that certain symptoms had a more significant impact due to the rigorous demands of their training and competitive environments. For instance, delayed reaction time proved particularly detrimental during critical moments in competitions requiring quick reflexes. Maintaining focus over extended periods was notably challenging, and exertion levels exacerbated feelings of fatigue, affecting performance. Additionally, severe muscle cramps were a common issue that often required immediate attention to prevent complications during competitions.

#### Stress-Induced Hyperglycemia

Athletes reported a relationship between stress and hyperglycemia, particularly before and during competitions. A participant stated, “Before [competition], I have so much adrenaline that my BG skyrockets,” and during competition, “No matter what I do, the adrenaline gets going, and I could start at a perfect six and feel great. And then as the [game starts], you can see my numbers go up to 20 to 25.” These descriptions suggest that anticipatory stress and competitive environments contribute to acute BG elevations. Additionally, a participant noted, “I’m stressed because I see my BG not being optimal […], and then I’ll be even more stressed […] So, I’m more reactive than proactive.” This demonstrates the vicious cycle that the athletes reported between BG levels and HRA.

#### Hyperglycemia-Related Anxiety

Beyond the physical symptoms, hyperglycemia led to constant stress that affected the athletes’ mental well-being, leading to HRA, “On a more psychological level, it’s a constant stress if [my BG] is not perfect or at the number I would want it to be for optimal performance.” The chronic nature of this stress was described as “in the background of your mind all the time.”

Additionally, fear of underperforming due to hyperglycemia added another layer of psychological strain, “In hyperglycemia, I don’t feel good. I don’t want to feel unwell during my sport and perform poorly or make mistakes that I wouldn’t normally make.” Feeling uncomfortable discussing it with teammates who cannot relate worsened the psychological impact for many, “You can’t really talk to a teammate about [HRA]. It’s not like it’s a pulled groin or a torn ligament that you can relate to most other people about.” Athletes expressed how stress affects their BG variation, emphasizing the need for targeted strategies. These strategies should aim to manage stress, thereby improving glycemic management and overall performance.

### Strategies Currently Used to Prevent HRA

Elite athletes discussed their strategies for reducing hyperglycemia during competition, therefore limiting HRA.

#### 1. Insulin and Nutrition

Athletes mentioned increasing basal insulin before a competition to prevent hyperglycemia. A participant stated, “I went from taking five units to eight units leading into that [competition], which is pretty substantial” to ensure normalized BG levels. Athletes also discussed increasing bolus insulin mid-game, as pre-competition adjustments were insufficient, highlighting the challenges of predicting insulin needs during competition, “I take more insulin because sometimes [my BG] continues to rise [mid-competition].”

Athletes stressed the importance of pre-planning meals and maintaining dietary consistency while being mindful of the glycemic index of foods. Many opted for complex carbohydrates to prevent rapid elevations; others focus on the timing and order of nutrient intake to control digestion rates, “Eating protein before carbs puts a blanket around your carbs, so you’re not eating carbs on an empty stomach and digesting them on their own. Having a little bit more fat with it also slows down the digestion rate. You’re not getting that [BG] spike all at one time.” Athletes also emphasized the importance of staying well-hydrated, highlighting its impact on mitigating their BG variability.

#### 2. Social Support Network

Engaging with healthcare professionals (HCPs) was highlighted as crucial for minimizing HRA, “My family doctor makes appointments with me every two weeks and we go through all my [rtCGM] data […] and if there’s a time where my BG isn’t ideal, it’s like, ‘Hey, I really need to dial in […] maybe instead of doing this, you need to do this.” The involvement of endocrinologists was also discussed, with a participant sharing that their endocrinologist provided them with potential strategies based on another of their patients’ experiences, “My endocrinologist is [a professional athlete’s] endocrinologist. […] I know that on the bench, he has a tablet that is directly connected to his sensor. As soon as he comes off the ice, he sees his BG automatically. If it’s rising, falling, he can control everything; it’s all at his fingertips.” Athletes described the importance of consultations with trusted HCPs to optimize diabetes management.

Furthermore, the study highlighted the support of dietitians and mental performance coaches. Dietitians played an essential role in guiding dietary choices and offered insights into managing BG through strategic meal planning. Mental performance coaches contributed to psychologically managing diabetes, particularly in navigating competitive stress challenges. None of the athletes mentioned that their training staff provided assistance or had an adequate understanding of T1D.

Family and friends were also recognized as crucial support systems, providing emotional support and practical assistance in diabetes management, “My parents are a good outlet. […] They get all my CGM data on their phone. If I see [my mom] text me at like […] 11:00 pm and she’s probably half asleep, I’m thinking, ‘I got to check my BG.’” These support systems contributed significantly to the athletes’ well-being, thereby reducing HRA.

Additionally, peer support emerged as a valuable strategy, “One of my friends has T1D and he’s a lifesaver, helped me a lot. He gives me some tips.” The exchange of insights among T1D peers provided unique support, as athletes drew on shared experiences to gain practical advice and emotional encouragement.

The study also acknowledged that following and interacting with influencers, celebrities, and professional athletes, who share their T1D experiences on social media helped to reduce HRA, “[A professional athlete] also has T1D, and he posts stuff on Instagram, so […] sometimes I think, ‘Oh, I could try that.’”

#### 3. Technology

Athletes unanimously highlighted rtCGMs’ importance in improving their BG level awareness and adjustments during competitions. They stated, “It can help during competitions to predict how my [BG levels] will go” and “Constantly having [rtCGM] at my fingertips, it can tell me exactly what I’m at and if I need to make any adjustments.” Two athletes mentioned that integrating rtCGM alerts into their daily routines was a crucial tool for preventive adjustments, allowing them to address BG variations before they become problematic.

Athletes highlighted the benefits of CSII for adjusting basal rates and making real-time adjustments, especially during training and competition, “I use a [CSII], and it really helps me […]. If you’re able to alleviate some decisions, that to me is very impactful to my psychological and physical well-being.” Furthermore, the integration of rtCGM with CSII technology emerged as a notably effective diabetes management approach. This combination addresses the dynamic interaction between CSII and rtCGM, offering athletes enhanced control and flexibility in managing their diabetes during various activities.

#### 4. Psychological Preparation

Athletes highlighted the role of meditation in managing HRA, “I try to focus a lot on the mental side of meditating pre-game to avoid that huge adrenaline spike.” They specifically mentioned meditation as crucial for clearing the mind the night before their competition, “Meditation is a big thing. I always meditate the night before the [competition]. It helps relax my nervous system because there’s a lot of anxiety about [competing].”

Additionally, athletes described the positive effects of mental preparation on BG stability, “I did mental preparation this year for the first time, and it was really cool. Stress raises your BG, and this year, […] my BGs were much more stable.” This effect suggested a potential link between mental preparedness, reduced stress, and improved BG levels. They also stated, “Being more prepared, being more in control, being more comfortable – obviously, I’m less stressed, so it’s sure that my BG rises less.”

Athletes also highlighted the efficacy of goal setting in mitigating HRA. However, they cautioned against having too many goals, as it can intensify anxiety and stress, “When you have 14 [goals] coming at you at once, it can seem overwhelming and amplify anxiety. When it’s just one, two or three [goals], it’s a little bit easier.” Additionally, the importance of setting realistic goals was mentioned, “If you set unrealistic goals […], it’s really easy to get let down.”

Finally, athletes noted the importance of focusing on performance and avoiding additional stress, “I concentrate on my performance and kind of forget about diabetes during that time.” This performance-centric mindfulness approach helps them manage BG levels more effectively. Having self-awareness about the current circumstances is also essential to mitigate anxiety, “I’m high, I’m dealing with it, and trusting the insulin […] I will come down […] I can’t do anything more just now other than focus on my game.”

#### 5. Routines

On competition days, athletes emphasized implementing pre-established routines to predict BG variability better, “Having a game plan, an established routine before a game helps me and I try to stick to it.” This athlete also details their game-day routine, “On game days, I have a pre-established routine. I […] always have the same breakfast and make the same smoothie with the same quantities […] my goal is to minimize variability and create a sense of predictability in managing my BG levels,” which effectively reduced their HRA.

#### 6. Aerobic Exercise

Athletes highlighted the significance of engaging in aerobic exercise before competition, as a valuable HRA management tool. One athlete mentioned incorporating long walks after meals and before a competition, perceiving a potential decrease in stress and subsequently BG levels, “If you’re high, [it helps to] do a prolonged aerobic warm up. That’ll probably bring you into range.”

Another athlete timed their workouts with periods in which they typically had lower BG levels, such as mornings. By aligning aerobic exercise routines with biological patterns, the athlete leverages their body’s natural responses to optimize BG control. The athlete stated that by knowing their “[BG] patterns, I try to work around them and use them to my advantage.”

#### 7. Sleep

Athletes noted that insufficient or unrestful sleep could impair performance, thereby increasing anxiety and contributing to BG elevations the following day, “If I don’t get enough sleep or I’m not properly rested, I find that [my BG] can spike.” Implementing structured bedtime routines to enhance sleep quality was also reported. They added, “I’ll have herbal tea before I jump in the shower […] then I’ll stretch and meditate so that my body just has that rhythm […] that’s been a big help for me.” Late-night schedules and deviations from regular sleep patterns were identified as factors contributing to BG management challenges.

Athletes reported using natural sleep aids such as melatonin and magnesium to increase relaxation and improve sleep quality. However, challenges were identified such as experiencing morning fogginess, which affected their performance during early-morning competitions, “Our [training] starts at 9:00 am, so I’m up at 5:30 to 6:00 am. The melatonin [..] has me in a fog through the morning. You want to feel sharp […], you want to feel ready, especially when [every second] matters.” This underscores the importance of considering the timing and potential side effects of sleep aids in the context of athletic schedules.

### Development of Strategies to Support Elite Athletes with HRA

#### 1. Social Support Network

Given that there are few elite athletes with T1D, building social support networks can be challenging. A participant stated, “If I knew another athlete who has really tight control, like [a professional athlete], I’m sure he doesn’t have these same problems as me because he has a team of medical professionals.” This statement demonstrates the need for support networks to provide T1D athletes with a platform for shared experiences and insights. Another participant highlighted the importance of creating, “A community where you can exchange […] with people who live the same reality as you […] And I think that’s something that might be missing.”

Additionally, providing family and friends with credible information may improve their support, “Your family members […] or your partner, they may not understand [TID].” The athlete added that developing support strategies for family and friends is essential to bridging existing gaps.

Further, teams and athletic organizations’ support staff should have access to targeted education to enhance their understanding of diabetes management, create an environment conducive to the successful integration of athletes with T1D, and reduce stigma. A participant stated, “It often happens that coaches ask me if I manage [my T1D] well. They don’t necessarily ask more questions. I think it comes from ignorance […] it’s not that they don’t care; it’s that they don’t know.” This comment underscores the need for education and training programs for coaching and support staff to ensure a comprehensive understanding of diabetes. They added, “it would help to know that the coaches are more aware of it. Maybe discussing it after a [competition] to know, ‘Okay, they understand that hyperglycemia had an impact on my performance,’ or ‘It’s a challenge in my sport performance.’” Athletes preferred training staff to understand T1D instead of taking an active role in their management, “The support that, someone like a trainer could provide would be having certain tools accessible [to understand T1D] rather than […] offering management tips.”

HCPs play an essential role in guiding athletes on diabetes management strategies and providing tailored advice. However, few HCPs have knowledge about the unique needs of elite athletes with T1D, let alone about HRA in this population.

#### 2. Technology

Many athletes desired systems that automate insulin adjustments based on factors like exercise and carbohydrate intake. These technologies have the potential to impact psychological and physical well-being significantly, “It’d be great to have a closed-loop system where I don’t have to put in how many carbs I’m eating. Or, like, if I’m working out, it’s just able to adjust my insulin values [without manual adjustments].” However, current technologies often fall short of meeting their expectations. Furthermore, the affordability of existing technologies remained a concern, “I find [rtCGMs] to be a technology that would be worthwhile. But you know, it’s expensive.”

Additionally, the discomfort and lack of trust associated with current CSII designs were highlighted. Some athletes refrained from using CSII due to concerns about the devices being large, bulky, and potentially unreliable, “I don’t feel comfortable playing with it, with wires […] I just wouldn’t trust it to not fall off.” Thus, redesigning CSIIs to meet elite athletes’ needs may make them more appealing.

Strategies are summarized in Figure 2.

**Figure 2.**
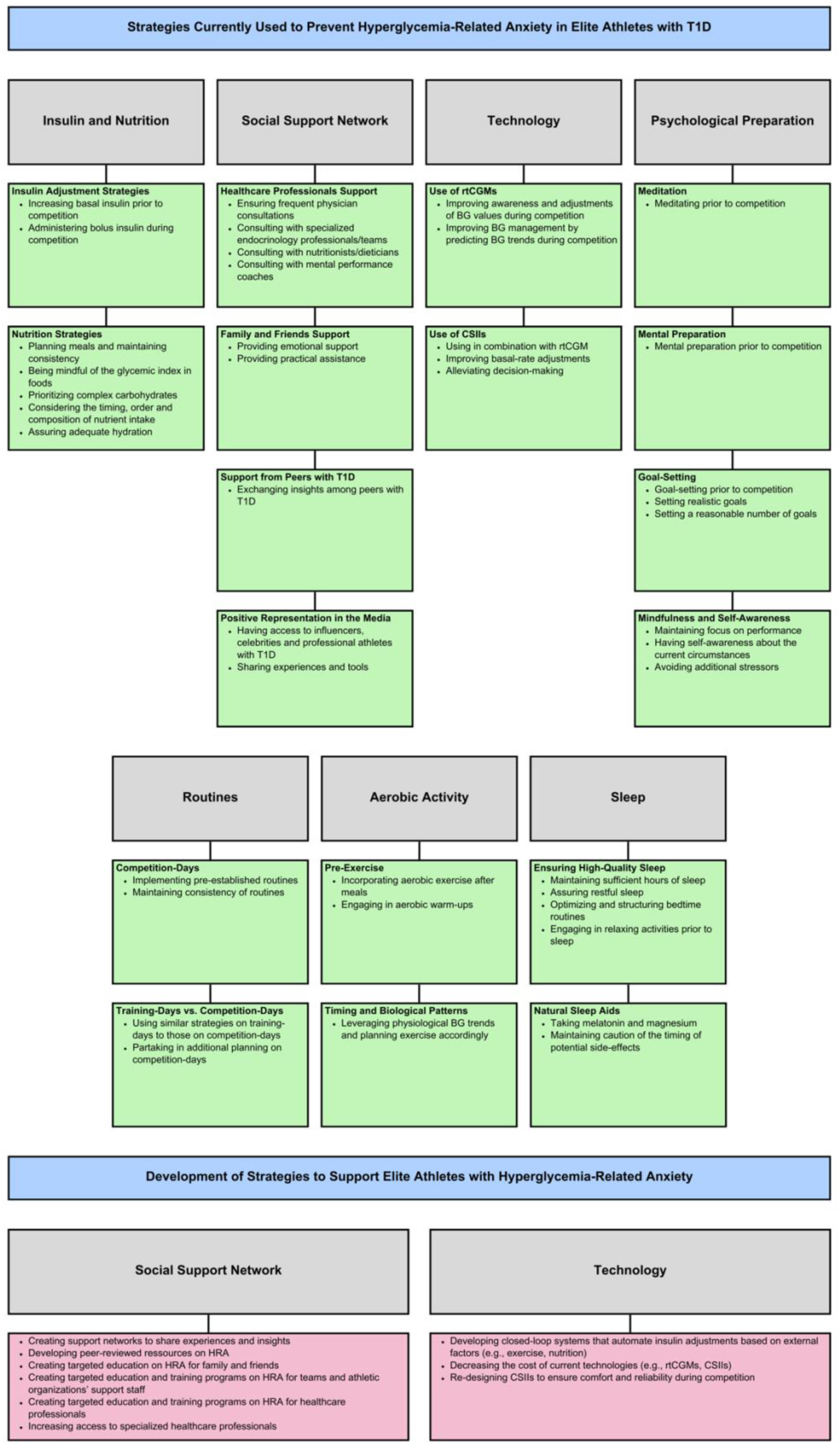
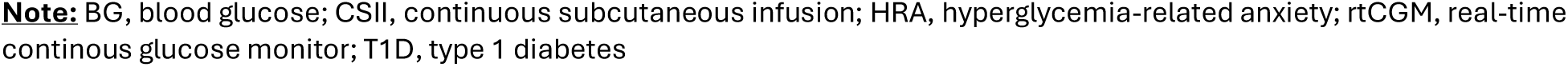
Current and Future Strategies to Support Elite Athletes with T1D and HRA

## Discussion

This qualitative study is the first to explore HRA in elite athletes with T1D. Athletes currently use numerous strategies to reduce HRA, however, they are often insufficient. A recurring theme was the critical role of maintaining well-controlled BG levels to reduce HRA risk, but athletes highlighted the impact of competition on hyperglycemia due to competition-related physical and emotional stress likely increasing stress hormones and glycemia [26–30]. While guidelines recommend checking BG frequently to understand its direction and magnitude of change [12, 15], this strategy is not always easy to implement during competition.

Consistent with previous findings [14, 30], trial-and-error approaches were prevalent among athletes with T1D. The limited documentation of tactics leaves elite athletes with few strategies [14] and a desire for additional resources. These resources could include peer mentoring, support from training staff, and the use of medical devices [29]. Moreover, athletes expressed challenges associated with CSII usage during exercise, including physical interference with sporting activities, a sentiment expressed elsewhere [6].

In Riddell et al (2020) [6], a few strategies were outlined for competitive athletes with T1D who experience competition-related hyperglycemia. These strategies include cognitive techniques, pre-competition aerobic exercises, and administering temporary basal rate increases or partial bolus insulin to correct hyperglycemia. However, they did not address HRA, similar to other scientific literature. Further, our participants’ experiences align with the suggestion from the *National Athletic Trainers’ Association* [12] regarding consulting physicians for increased basal rates or small insulin boluses during periods of exercise-induced hyperglycemia. However, there are no evidence-based guidelines that physicians can use to guide their patients in this population. Thus, there is a need to develop resources to support elite athletes with T1D [15], particularly in subject areas such as HRA. Studies have shown that having access to specialized healthcare resources that are co-developed with patients can increase confidence in diabetes management [31, 32]. Thus, future efforts should focus on developing education and awareness programs tailored to the unique needs of elite athletes with T1D, addressing the psychological and physiological challenges associated with competition-related hyperglycemia. Future strategies should be adaptive, accommodating each athlete’s unique diabetes management needs [30].

Our study has several notable strengths and limitations. We implemented bracketing and member checking to increase reliability. The exclusion of participants with GAD limits this potential confounding variable. The broad spectrum of sports represented can be seen as a strength or a weakness; while it allowed us to gain a comprehensive understanding of HRA-related consequences and strategies, it limited the detailed exploration of specific challenges within each sport. Despite the small sample size of 10 participants, thematic saturation was achieved after approximately 5 or 6 interviews, with no new themes emerging in the subsequent interviews. This suggests that the sample size, while limited, was adequate for our qualitative analysis. Our study population, predominantly including Caucasian individuals from developed nations, may limit the generalizability of the results. Future research should consider larger, more diverse samples to enhance applicability.

## Conclusion

In conclusion, addressing HRA in elite athletes with T1D involves refining current strategies, developing personalized tools, bridging support system gaps, enhancing education for athletes, training staff, peers, and HCPs, and acknowledging the unique needs of each athlete in the context of competition. Future research should aim to fill the existing void in resources and support mechanisms to optimize T1D management and performance in this population.

## Data Availability

Data cannot be shared publicly as it contains information that can reveal participants identity. Please contact study researchers for more information.

## Acknowledgments

The authors would like to thank the BETTER study group for guidance in establishing the research question and designing the platform. The authors would also like to thank the participants, Erik Sesbreno and the patient partner for their contributions to this project.

## Funding and Assistance

This project was funded by Chaire J.A DeSève. A.K. received funding from the Canadian Graduate Scholarship-Master’s Program. AS.B. is a fond de recherche du Québec en santé research scholar. All other authors received no financial support for the research, authorship, and/or publication of this article.

## Author contributions and guarantor statement

A.K.: Conceptualization, methodology, project administration, protocol - original draft, protocol -review & editing, participation recruitment, interview conduction, formal analysis, visualization, writing - original draft, writing - review & editing. A.S.: Conceptualization, methodology, protocol - original draft, protocol - review & editing, interview conduction, formal analysis, visualization, writing - original draft, writing - review & editing. MA.F.: Interview conduction, writing - original draft, writing - review & editing. J.Y.: Conceptualization, protocol - review & editing, participation recruitment, writing - review & editing. J.K.: Conceptualization, methodology, protocol - review & editing, writing - review & editing. M.K.T: Protocol - review & editing, writing - review & editing. A.H.: Protocol - review & editing, writing - review & editing. R.R.L.: Conceptualization, protocol - review & editing, writing - review & editing, supervision. AS.B.: Conceptualization, protocol - review & editing, writing - review & editing, supervision. AS.B. is the guarantor of this work and, as such, has full access to all the data in the study and takes responsibility for the integrity of the data and the accuracy of the data analysis.

## Prior Presentation

Presented at the 84th American Diabetes Association conference in June 2024 in Orlando, FL, USA.

## Abbreviations

BETTER: Behaviours, Therapies, Technologies, and Hypoglycemia Risk in T1D
BG: Blood glucose
CSII: Continuous Subcutaneous Insulin Infusion
GAD: Generalized Anxiety Disorder
HCP: Healthcare Professional
HRA: Hyperglycemia-Related Anxiety
IRCM: Montreal Clinical Research Institute
MDI: Multiple Daily Injection
PA: Physical Activity
REDcap: Research Electronic Data Capture
rtCGM: Real-Time Continuous Glucose Monitors
T1D: Type 1 Diabetes

